# Pathogenic Risks in Courier-Based Food Delivery Systems: Integrating Microbiological Surveillance into Zambia’s Food Safety Framework

**DOI:** 10.64898/2026.04.04.26350179

**Authors:** Choongo Mulungu, Zimba Newstead, Lalisa Nambeye, Derby Samu, Muyembe Gladys, Chaldren Kaluah, Christine Musonda, Astridah Kona Yihemba Maseka

## Abstract

**Research background:** Foodborne diseases (FBDs) remain a pressing global public health issue, with courier-based food delivery systems increasingly recognized as potential contamination pathways. In Zambia, despite the Food Safety Act No. 7 of 2019, limited evidence exists on microbial risks in courier-mediated food transport. This study was conducted to assess pathogenic contamination in food carriers used by courier bikers in Lusaka during the 2025/2026 cholera outbreak response.

**Experimental approach:** An analytical cross-sectional design was employed. Ninety-three food carriers (bags, cooler boxes, and metal containers) were randomly sampled from courier bikers. Swabs from internal surfaces were processed within 24 hours using standard microbiological culture and biochemical identification methods. Statistical analyses (chi-square tests, Pearson correlations, and logistic regression) were applied to determine associations between contamination and operational factors.

**Results and conclusions:** Microbial contamination was detected in 69% of carriers. The most common pathogens were Escherichia coli (30%), coagulase-negative Staphylococcus (24%), and Staphylococcus aureus (18%), with additional isolates including Gram-positive bacilli (11%) and Klebsiella pneumoniae (8%). Logistic regression identified cleaning frequency as the strongest predictor of contamination, with infrequent cleaning associated with significantly higher odds ratios (26.5–94.7, p < .05). Carrier type also influenced contamination risk, while years in service and certification status were not significant. The findings highlight that inadequate cleaning practices and carrier design are primary drivers of microbiological risks in courier-based food delivery systems.

**Novelty and scientific contribution:** This study provides the first empirical evidence of microbial contamination in courier food carriers in Lusaka, Zambia. It underscores the urgent need for strengthened hygiene protocols and routine sanitation enforcement to protect consumers from foodborne pathogens and antimicrobial resistance. The work contributes novel insights into food safety risks in emerging delivery systems, with implications for policy, public health interventions, and consumer protection in Zambia and beyond.

## INTRODUCTION

Food safety remains a pressing global public health concern, with foodborne diseases (FBDs) contributing significantly to morbidity and mortality worldwide [*1,2*]. The World Health Organization (WHO) estimates that unsafe food causes over 600 million cases of illness annually, with pathogens such as *Salmonella spp., Escherichia coli, Listeria monocytogenes, Campylobacter spp*., and *Staphylococcus aureus* among the most prevalent etiological agents [*3,4*]. Beyond microbial hazards, chemical contaminants and poor handling practices exacerbate risks, underscoring the multifactorial nature of food safety challenges [*5*]. Comparable concerns are evident in Europe, where surveillance data reveal recurrent outbreaks linked to zoonotic pathogens and poor food handling. Between 2015 and 2019, major FBD outbreaks across the European Union highlighted persistent risks from Salmonella, Listeria, and Campylobacter [*6*]. The WHO European Region estimates that unsafe food contributes significantly to the burden of disease, with children under five disproportionately affected [*7*]. More recent analyses confirm a surge in outbreaks and fatalities between 2008 and 2022, emphasizing the need for integrated One Health approaches to mitigate emerging threats [*8,9*].

The African region bears a disproportionate burden of FBDs due to infrastructural limitations, weak enforcement of food safety regulations, and inadequate training among food handlers [*10,11*]. Studies in Ghana and other Sub-Saharan countries have documented unsafe courier and food handling practices, highlighting courier-based delivery systems as emerging contamination pathways [*11,12*]. Microbiological assessments of delivery boxes in Accra revealed contamination with pathogens such as *E. coli* and *Staphylococcus aureus*,emphasizing the vulnerability of last-mile delivery systems [*13*]. Similarly, research on street food vendors in Lusaka identified persistent hygiene gaps, reinforcing the need for systemic interventions [*12*].

In Zambia, microbial risks have been documented across traditional food chains. Phiri et al., [*14*] identified contamination in dairy value chains, while Mumbula et al., [*15*] reported microbial loads in edible insects. More recently, Mulungu et al., [*16*] highlighted gaps in food safety training programs for food handlers, pointing to systemic weaknesses in regulatory compliance. Despite the enactment of the Food safety Act No. 7 of 2019, which provides comprehensive provisions for food safety regulation and enforcement, courier-based food delivery systems remain largely undocumented in Zambia’s food safety literature. This is concerning given the rapid growth of online food delivery in Lusaka, where motorcycles and bicycles are increasingly used to transport ready-to-eat meals.

Microbiological surveillance of food carriers is critical, as studies elsewhere have demonstrated contamination risks in reusable plastic bags [*17*] and cold chain packaging systems [*18,19*]. In Zambia, where courier-based food delivery is expanding rapidly, the absence of systematic microbial risk assessments represents a critical gap in safeguarding consumers against FBDs.

## MATERIALS AND METHODS

### Study design

An analytical cross-sectional quantitative study was conducted to assess microbial contamination in food carriers used by courier bikers in Lusaka, Zambia. The study focused on microbiological sampling and analysis to provide direct evidence of contamination risks in courier-based food delivery systems.

### Procedure

Courier bikers actively engaged in food delivery within Lusaka were approached at bike ranks, delivery hubs, shopping malls, markets and streets. Inclusion criteria required participants to be aged 18 years or older, actively delivering food, and willing to allow for swabbing for microbiological analysis. A total of 93 food carriers including thermal bags, cooler boxes, Fiber boxes, plastic buckets, metal boxes and food delivery bags were sampled using simple random sampling. This sample was larger than prior research by Budhathoki et al., [*20*] which included only 34 carriers. Participants who declined were replaced by the next randomly selected biker until the target size was reached. Microbial saturation was achieved by the 46^th^ swab, with no new organisms isolated thereafter. Extending to 93 carriers confirmed saturation and strengthened reliability without unnecessary duplication. Given the concurrent cholera outbreak, resources limited further analysis beyond doubling the saturation point. This approach aligns with international food safety studies emphasizing saturation in environmental sampling [*17,13*].

Swab samples were collected from the internal surfaces of food carriers using sterile cotton swabs moistened with buffered peptone water. Each swab was applied to a 25 cm^2^ area of the carrier surface, following standardized protocols for environmental sampling in food safety research [*3,17*]. Samples were immediately placed in sterile transport tubes, labeled, and maintained under cold chain conditions during transportation to microbiology laboratories in Lusaka.

### Microbiological analysis

Samples were processed within 24 hours of collection. Standard culture methods were employed to isolate and identify microorganisms commonly implicated in FBDs, including *Salmonella spp., E. coli, Listeria monocytogenes, Campylobacter spp., Staphylococcus aureus*, and *Clostridium perfringens* [*3,17*].

Microbial identification was performed using selective and differential media, followed by biochemical testing. Colony counts were expressed as prevalence percentages of positive samples. Quality assurance procedures included duplicate plating and the use of reference strains for validation.

### Statistical analysis

Microbiological data were analyzed descriptively, reporting the prevalence and distribution of isolated pathogens across the 93 food carriers. Statistical analysis was conducted using JASP 0.19.0.0, with chi-square tests applied to examine associations between contamination and selected courier characteristics. Pearson coefficient correlation was employed to determine the relationships between variables while Logistic regression analysis was performed to estimate the predictors of contamination in the sample.

## RESULTS AND DISCUSSION

### Sociodemographic and operational characteristics of courier bikers

**Table 1**, the study included 93 courier bikers with a mean age of 28.9 years (range = 18–48). All participants were males, the majority of participants were educated to the secondary level (75.3%) and operated as independent contractors (58.1%). Regarding equipment, most of the participants utilized motorbikes (78.5%) and food bags as their primary carrier type (66.7%). Notably, only 24.7% of the bikers were certified food handlers, and a combined 81% had been in service for at least one year.

**Table 1:** Sociodemographic and operational characteristics of courier bikers (N = 93)

| Variable | Categories | Frequency (n) | Percentage (%) |
| --- | --- | --- | --- |
| Age (years) | Mean = 28.9, Range = 18–48 | n.d | n.d |
| Education | No school | 3 | 3% |
|  | Primary | 12 | 13% |
|  | Secondary | 70 | 75% |
|  | Tertiary | 8 | 9% |
| Certified Food Handler | Yes | 23 | 25% |
|  | No | 70 | 75% |
| Mode of Operation | Employed | 39 | 42% |
|  | Independent | 54 | 58% |
| Years in Service | <1 year | 18 | 19% |
|  | 1–2 years | 38 | 41% |
|  | 3–5 years | 37 | 40% |
| Bike Type | Motor Bike | 73 | 78% |
|  | Bicycle | 20 | 22% |
| Carrier Type | Food Bag | 62 | 67% |
|  | Bucket | 3 | 3% |
|  | Metal Box | 12 | 13% |
|  | Cool Box | 12 | 13% |
|  | Fiber Box | 4 | 4% |
| Contaminated carrier | Yes | 64 | 69% |
|  | No | 29 | 31% |

n.d = not detected, n = frequency in number

### Pathogen prevalence and hygiene associations among courier biker carriers

**Table 2**, details microbial analysis of the courier carriers revealed a high prevalence of contamination. The most frequently isolated pathogen was E. coli (*n* = 28, 30%), followed by Coagulase-negative Staphylococcus (*n* = 22, 24%),) and Staphylococcus aureus (*n* = 17, 18%),). Notably, 26% (*n* = 24) of the carriers tested positive for more than one organism.

**Table 2.**
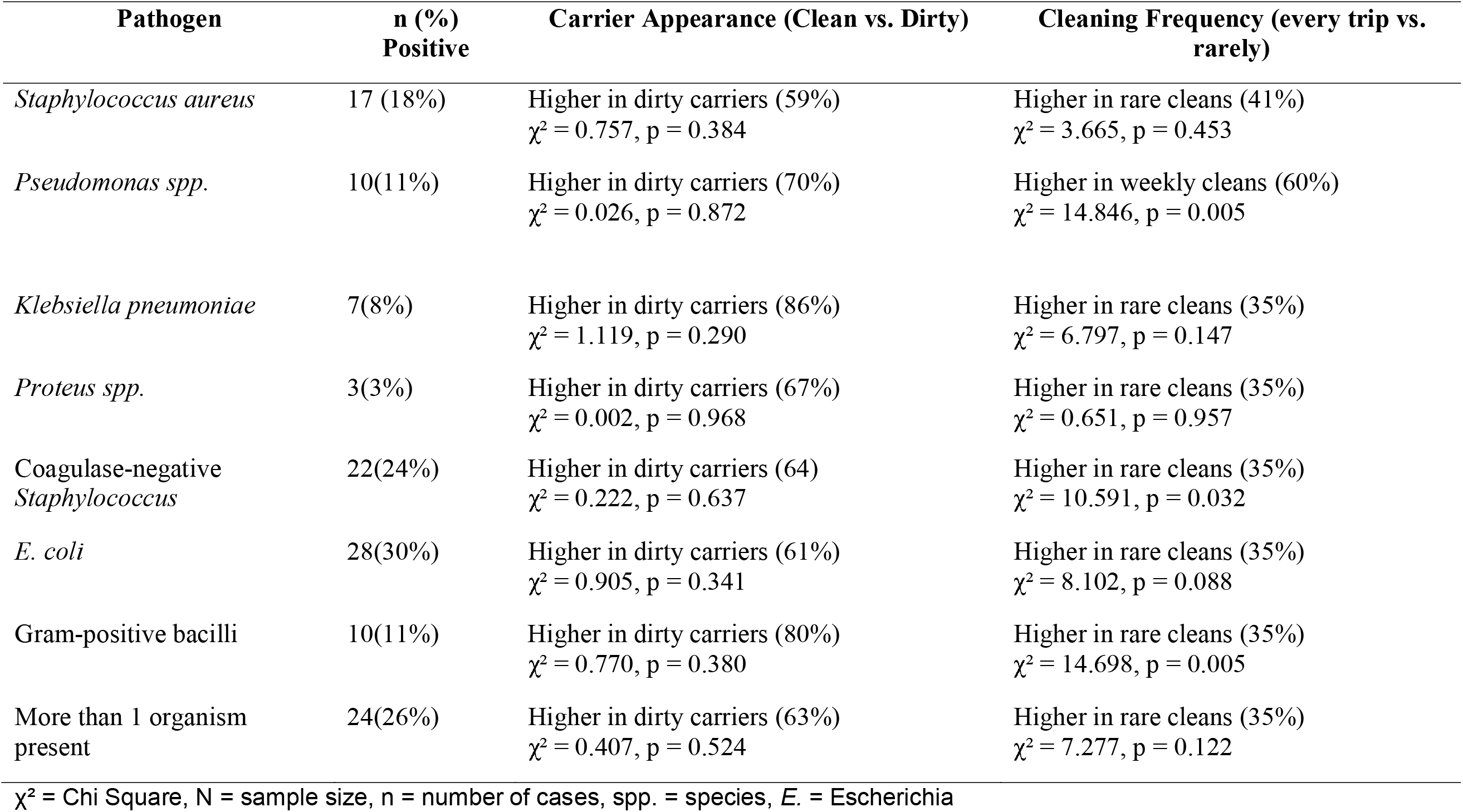
Pathogen prevalence and hygiene associations among courier biker carriers (n = 93)

Chi-square tests were conducted to determine if carrier appearance or cleaning frequency were associated with the presence of pathogens. Interestingly, visible carrier appearance (clean vs. dirty) did not significantly associate with the presence of any specific pathogen (all *p* > .05). While pathogens were consistently “higher in dirty carriers” by percentage, visible cleanliness was not a statistically significant predictor of microbial safety. However, cleaning frequency showed several significant associations. The presence of Pseudomonas spp. was significantly associated with cleaning frequency, χ^2^ (1) =14.85, *p* = .005, with higher rates found in carriers cleaned weekly. Significant associations were also found for Coagulase-negative Staphylococcus (χ^2^ (1) = 10.59, *p* = .03) and Gram-positive bacilli (χ^2^ (1) = 14.70, *p*= .005), both of which were more prevalent in carriers that were rarely cleaned. No significant association was found between cleaning frequency and the presence of E. coli (*p*= .088) or S. aureus (*p* = .452).

### Relationships between Carrier Contamination Classification and other Variables

A Pearson’s correlation analysis was conducted to examine the relationships between carrier contamination and various operational factors (**Table 3**). Carrier classification (or contamination status) was found to have a significant positive correlation with frequency of cleaning (*r* = .279, *p* = .007), carrier type (*r* = .268, *p* = .009), and bike type (*r* = .229, *p* = .027). While certified food handlers were significantly more likely to maintain a carrier with a clean appearance (*r* = .351, *p* = .001), neither certification nor physical appearance showed a significant linear relationship with the actual microbiological contamination status of the carrier (*p* >.05). Additionally, a strong positive correlation was observed between carrier type and frequency of cleaning (*r* = .378, *p* = .001), suggesting that cleaning behaviors vary significantly across different container materials. Results indicate a significant negative correlation between Bike type and appearance of the carrier (*r*= -.249, *p* = .016) suggesting that motor bikers were associated with cleaner carriers. Years in service is positively correlated with appearance of the carrier (*r* = .220, *p* = .034) suggesting that bikers with more experience tended to have cleaner carriers while age of the biker was positively correlated with the frequency of cleaning (*r* = .211, *p* = .042) implying that age increased with increase in frequency of cleaning their carriers.

**Table 3:** Relationships between carrier contamination classification and other variables.

| Variable | 1 | 2 | 3 | 4 | 5 | 6 | 7 | 8 |
| --- | --- | --- | --- | --- | --- | --- | --- | --- |
| 1. Carrier Classification | — |  |  |  |  |  |  |  |
| 2. Certified Food Handler | -.004 | — |  |  |  |  |  |  |
| 3. Bike Type | .229* | -.057 | — |  |  |  |  |  |
| 4. Years in Service | -.102 | -.005 | .090 | — |  |  |  |  |
| 5. Carrier type | .268** | .163 | -.040 | .175 | — |  |  |  |
| 6. Appearance of Carrier | -.049 | .351*** | -.249* | .220* | -0.062 | — |  |  |
| 7. Frequency of Cleaning | .279** | .041 | .155 | -.147 | .378*** | -.201 | — |  |
| 8. Age of Biker | .119 | .145 | .104 | -.165 | .136 | -.101 | 0.211* | — |

\* p < .05, \*\* p < .01, \*\*\* p < .001

### Regression analysis

The logistic regression model in (Table 4) examined years in service, carrier type, and frequency of cleaning as predictors of contamination. As shown in Table 5, years in service was not a significant predictor (*B* = 0.054, *SE* = 0.230, OR = 1.056, 95% CI [0.67, 1.66], *p* = .814), indicating that experience did not reliably reduce contamination risk. In contrast, carrier type (1) was significantly associated with lower odds of contamination (*B* =-5.224, *SE* = 2.470, *OR* = 0.005, 95% CI [0.01, 0.68], *p* = .034), suggesting that certain carrier designs may offer protective effects. Finally, frequency of cleaning emerged as a strong and consistent predictor: carriers cleaned less frequently had markedly higher odds of contamination, with odds ratios ranging from 26.5 to 94.7 across categories (all p < .05). These findings highlight that operational hygiene practices, particularly cleaning frequency and carrier type, are more influential determinants of contamination risk than years of service. Overall, the model combining all predictors was highly significant (Δχ^2^ = 42.98, p <.001), with Nagelkerke R^2^ = 0.493, indicating that the set of operational and demographic factors explained nearly half of the variance in contamination risk.

**Table 4:**
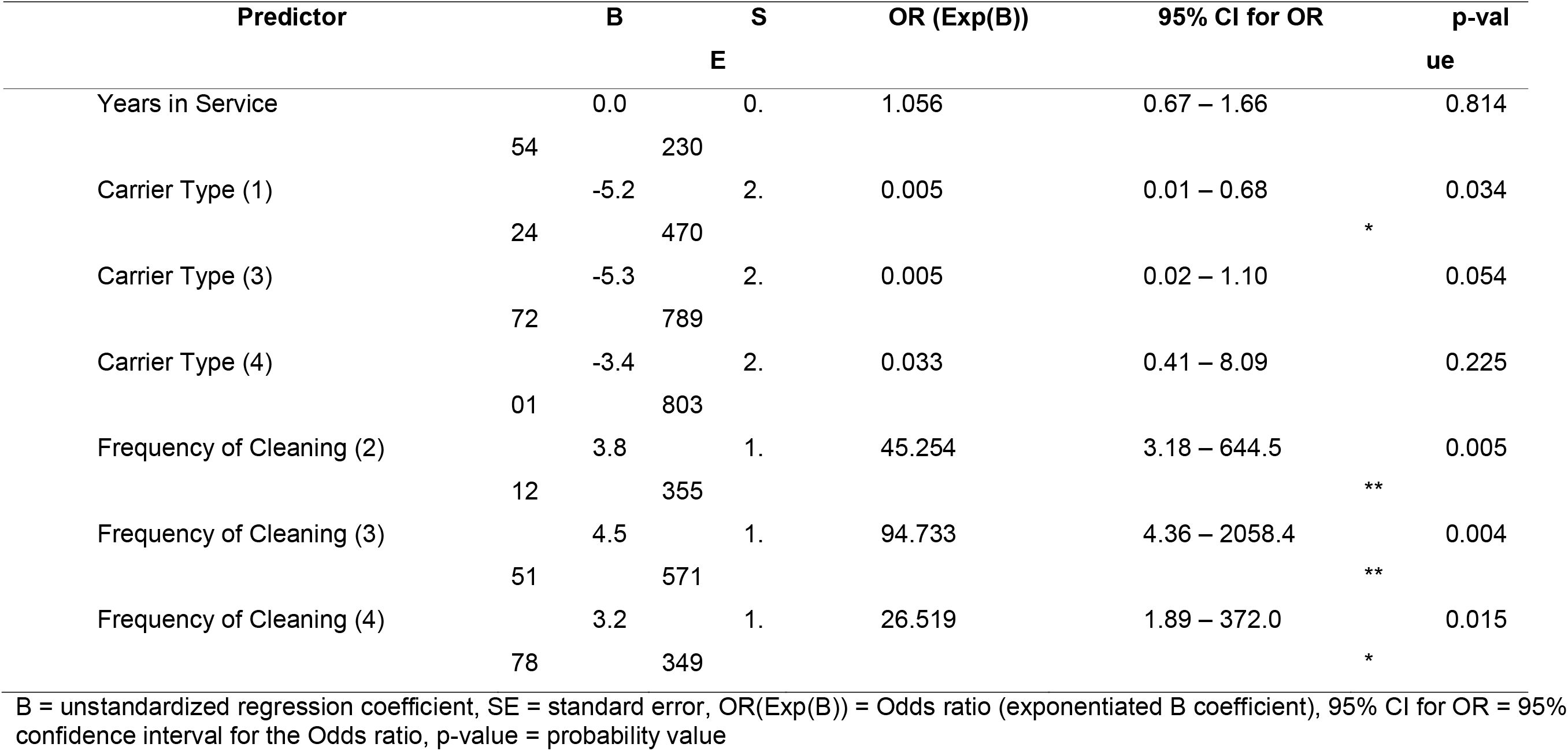
Regression analysis (logistic regression – odds of contamination)

## Discussion

This study revealed a substantial prevalence (69%) of microbial contamination among courier biker carriers, with *Escherichia coli* (30%), coagulase-negative *Staphylococcus* (24%), and *Staphylococcus aureus* (18%) most frequently isolated. Notably, 26% of carriers harbored more than one organism, underscoring the compounded risk of cross-contamination during food transport.

The presence of Gram-negative bacilli such as *E. coli* and *Klebsiella pneumoniae* aligns with findings from hospital and food chain studies, where these organisms were frequently isolated from equipment and surfaces [*21-23*]. *Klebsiella pneumoniae*, in particular, has been increasingly recognized as an emerging foodborne hazard, often carrying antibiotic resistance traits [*24,23*]. These findings highlight the concern that courier-mediated food distribution may contribute to the spread of resistant pathogens.

Gram-positive bacilli, including *Bacillus cereus*, are well-documented causes of food poisoning, particularly in ready-to-eat meals [*25,26*]. Their presence in carriers that were rarely cleaned suggests that inadequate hygiene practices facilitate spore persistence and toxin production, increasing outbreak risk. Similarly, coagulase-negative *staphylococci* (CoNS), though less pathogenic than *S. aureus*, possess enterotoxigenic potential and were significantly associated with carriers cleaned infrequently [*27,17*].

Importantly, visible carrier cleanliness was not a reliable predictor of microbial safety. Pathogens were more common in carriers classified as “dirty,” but statistical analysis showed that cleaning frequency was the strongest determinant of contamination risk, with odds ratios as high as 94.7 for carriers cleaned infrequently. This emphasizes that operational hygiene practices, rather than superficial appearance, are critical for food safety.

Carrier type also influenced contamination risk, with certain designs associated with lower odds of contamination. This supports evidence that material and structural properties affect microbial persistence and cleaning behavior [*18,19*].

Sociodemographic factors such as years in service and certification status did not significantly predict contamination, although certified food handlers were more likely to maintain carriers with a clean appearance. This parallels findings in Zimbabwe and Zambia, where outbreaks were linked more to systemic hygiene practices than to individual training [*28,14*]. Consumer concerns about hygiene in motorcycle food delivery services further reinforce the need for stronger sanitation protocols [*29*.*30*].

### Limitations

While this study provides valuable insights, several limitations should be acknowledged. First, the sample size was limited to 93 courier bikers, which may restrict generalizability across broader delivery networks. Second, the study focused on bacterial contamination, leaving fungal and viral pathogens unexplored, despite evidence of fungal contamination in transport systems [*31*]. Third, the cross-sectional design precludes causal inference, and longitudinal studies would be needed to assess contamination dynamics over time. Environmental factors such as temperature fluctuations during delivery, which have been shown to influence microbial survival [*19*], were not directly measured. Finally, this study did not involve females due to none availability of females in the business. However, futures studies should endeavor to balance gender to identify any differences across gender.

## CONCLUSIONS

This study demonstrates that courier-mediated food delivery systems can act as significant vectors for foodborne pathogens, with contamination strongly influenced by cleaning frequency and carrier design. The identification of pathogens with established food poisoning potential highlights a critical gap in food safety oversight within this rapidly expanding sector. By providing the first empirical evidence from Lusaka, the findings underscore the novelty and urgency of addressing hygiene risks in courier-based food transport. Strengthening sanitation protocols, improving carrier design, and raising consumer awareness are essential steps to mitigate microbial hazards and curb the spread of antimicrobial resistance. These insights contribute to advancing food safety policy and practice, offering a timely scientific contribution with direct relevance to public health protection.

## Data Availability

All data produced in the present study are available upon reasonable request to the authors

## ACKNOWLEDGEMENTS

We thank the Lusaka District Health Office for logistical support to conduct the activity culminating into this paper at such a critical time. Further, we acknowledge the food courier bikers without whom this study wouldn’t have been possible.

We also thank the local authorities for the roles they played in ensuring this study is done in accordance with established protocols and their permission for us to conduct the study in their jurisdiction. We also acknowledge Mr. Realman Machalo and Mr. Mthunzi Banda for the roles they played during data collection.

## ETHICS APPROVAL

Ethical approval for this study was waived, as the investigation formed part of the official cholera outbreak response mandated under Ministry of Health protocols in Zambia. All procedures were conducted in accordance with national public health guidelines. Despite the waiver, informed consent was obtained from all participating courier bikers prior to sample collection, ensuring voluntary participation and respect for individual autonomy.

## AUTHORS’ CONTRIBUTION

C. Mulungu served as the principal investigator, conceptualized the study, and prepared the initial draft of the manuscript. Z. Newstead, an Environmental Health expert, together with L. Nambeye and A. K. Y. Maseka, both Public Health experts, and C. Musonda D. Samu, and C. Kaluah Laboratory experts, contributed as co-investigators and provided technical input. L. Nambeye, M. Gladys, and A. K. Y. Maseka facilitated access to local resources that supported efficient data collection. C. Mulungu, and Z. Newstead carried out field data collection with other assistants acknowledged in this paper, while C. Mulungu, C. Musonda, D. Samu, N. Zimba and C. Kaluah coordinated laboratory investigations and contributed to drafting the manuscript. All investigators critically reviewed the manuscript, strengthened the reporting of findings, and approved the final version for publication.

